# Status of hypertension control in urban slums of Central India : A Community health worker based two-year follow up

**DOI:** 10.1101/2021.02.02.21251036

**Authors:** Pakhare Abhijit, Lahiri Anuja, Shrivastava Neelesh, N Subba K, Veera Durga Vaishnavi Kurra, Joshi Ankur, Atal Shubham, Khadanga Sagar, Joshi Rajnish

**Author notes:** **Corresponding Author:** Name: Rajnish Joshi, Address: Baghsewania, AIIMS campus, Saket Nagar, Bhopal (462020), MP.

## Abstract

**Background:** Hypertension is a leading cause of cardiovascular diseases and its control is poor. There is heterogeneity in levels of blood-pressure control among various population sub-groups. The present study was conducted within the framework of National Program for prevention and control of cancer, diabetes, cardiovascular diseases and stroke (NPCDCS) in India. It aims to estimate proportion of optimal blood pressure control and identify potential risk factors pertaining to uncontrolled hypertension consequent to initial screening.

**Methods:** We assembled a cohort of individuals with hypertension confirmed in a baseline screening in sixteen urban slum clusters of Bhopal (2017-2018). Sixteen Accredited Social Health Activists (ASHAs) were trained from within these slums. Individuals with hypertension were linked to primary care providers and followed-up for next two years. Obtaining optimal blood-pressure control (defined as SBP< 140 and DBP<90 mm of Hg) was a key outcome.

**Results:** Of a total of 6174 individuals, 1571 (25.4%) had hypertension, of which 813 were previously known and 758 were newly detected during baseline survey. Two year follow up was completed for 1177 (74.9%). Blood-pressure was optimally controlled in 301 (26%) at baseline, and in 442 (38%) individuals at two years (absolute increase of 12%; 95% CI 10.2-13.9). Older age, physical-inactivity, higher BMI and newly diagnosed hypertension were significantly associated with uncontrolled blood-pressure.

**Conclusions:** We found about six of every ten individuals with hypertension were on-treatment, and about four were optimally controlled. These findings provide a benchmark for NPCDCS, in terms of achievable goals within short periods of follow-up.

## Introduction

Hypertension (HTN) is a leading risk factor of cardiovascular diseases and its control is poor. There occurs a loss of numbers at screening-awareness-treatment-control pathway. Among those with hypertension, in urban India less than half are aware of their elevated blood pressure, and only one in five individuals are controlled.(1) Studies from central India found that 22.3% of all individuals in the age group of 18-69 years had hypertension, and only 11.8% of those who had hypertension were being treated.(2) Better control of hypertension depends upon availability and access to quality health-care.(3) Once diagnosed, management of untreated and uncontrolled HTN is challenging. There needs to be a focus on improved adherence.(4) Various engagement strategies have been used, including but not limited to mobile-health applications, (5) telemedicine services, and community health worker (CHW) based engagement strategies.(6) While some population based studies have had a limited impact on blood-pressure levels, (7,8) other facility based studies have shown benefits.(9,10) Clusters where more members have uncontrolled hypertension at baseline, with a higher baseline blood-pressure, have reported greater and more significant reductions.(11) Even amongst a homogenous health-system, variation in blood pressure control exists, attributable to better patient engagement.(12) National Program for prevention and control of cancer, diabetes, cardiovascular diseases and stroke (NPCDCS) in India recommends annual hypertension screening for all adults, and proportion of optimally controlled individuals as a facility level indicator. Aim of the current study is to identify proportion of individuals with optimal blood pressure control and to identify risk-factors for uncontrolled hypertension.

## Methods

### Design and ethics

We established a community based cohort. The study design was approved by the Institutional Human Ethics Committee (Ref: IHEC-LOP/2017/EF00045) and funded by Indian Council of Medical Research. All participants provided written informed consent prior to initiation of any study procedures.

### Setting

The study was conducted in 16 urban slum clusters of Bhopal, a city with population of about 18 Lakhs, located in central India. A baseline door-to-door survey was conducted in the urban slum clusters between November 2017 and March 2018, details of which are provided elsewhere.(13) Briefly, all consenting non-pregnant adults above the age of 30 years formed a cross-section, in whom cardiovascular disease (CVD) risk assessment was performed by Accredited social health activists (ASHAs), who are CHWs in these communities. Study supervisors provided oversight, and also confirmed key variables, before stratifying individuals as high risk. Individuals identified at a high CVD risk, especially those with hypertension and diabetes were sought to be linked to urban primary health centres (UPHCs) which are primary-care public-health facilities in these communities. All participants were however free to seek care from alternate private-providers of their choice. Limited generic anti-hypertensive drugs (Amlodipine, Losartan, and Hydrochlorothiazide) are available at no-cost from UPHCs with a prescription refill duration varying from two to four weeks. Participants who visit private primary-care providers usually incur out-of-pocket expenditure for their drug therapy, with a refill duration dependent on their capacity to pay.

### Participants

All individuals residing in selected urban slum clusters of age 30 years or more were screened for hypertension as part of baseline assessment. Individuals with hypertension (either a previously known or a newly detected hypertension), were identified in a baseline survey. Individuals who reported themselves as diagnosed to have hypertension in the past, were defined as having *previously known-hypertension*. ASHAs obtained a first-set of blood-pressure values using a digital sphygmomanometer (Omron digital apparatus, Model 7200, Kyoto, Japan) at home of the participants. All the blood pressure readings were then confirmed in a second set of readings obtained by the study supervisor, using the same device. Each set of blood-pressure readings consisted of an average of three values, obtained one-minute apart, using a standardized technique. A person was defined as a *newly detected hypertensive*, if the blood pressure values were above Systolic blood pressure (SBP) of 140 mm Hg or a Diastolic blood pressure (DBP) of 90mm Hg on both the occasions. This is the definition for diagnosis of HTN under NPCDCS. All individuals with hypertension (previously known or newly detected) were provided with referrals for physicians available at UPHC, who initiated them with or optimized their anti-hypertensive medication. All participants, who completed the two-year follow-up assessment in November-December 2019 are included in the current study.

### Procedures

We followed individuals with hypertension (previously known or newly detected) for a period of two years. Follow-up was performed using the existing human resources in the public health-delivery system, facilitated by cloud-based data-management tools, and study supervisors. We prepared a list of all individuals with HTN in a database, and stratified them by cluster. We provided these lists of each cluster to respective ASHAs, who were responsible for visiting each member at-least once in every three-months. Purpose of these visits was to promote adherence to drug therapies, measurement of blood pressure, and to provide focussed CVD-prevention health education. Monetary incentives were given to ASHAs for every completed follow-up (rupees 50 INR about one USD). In each visit, ASHAs recorded linkage to UPHC or a private care provider, and continuation of drug therapy. We defined *treatment interruption* as failure to obtain a prescription-refill for two-weeks or more after exhaustion of previously obtained drug supplies.

We designed a cloud-based digital tool using CommCare based application to facilitate follow-up. This tool was available on mobile phones of all ASHAs, and in addition to facilitate follow-up, it provided access to baseline values of the participants, and also was to be used for data-entry of each subsequent visit. Study supervisors monitored cohort follow-up using this CommCare based application, and performed additional site-visits for troubleshooting. We also conducted a quarterly-meeting of all ASHAs and study supervisors to monitor follow-ups. A community clinic based one-year adherence assessment was performed by a team of pharmacologists who were trained to assess pill-burden and adherence to previously prescribed therapies. This assessment was limited to those who were on any anti-hypertensive drug therapy in the previous month.

### Outcome assessment

Blood pressure control at two-year assessment visit was the key outcome. It was defined as *optimally controlled* if average SBP was less than 140mm Hg and DBP was less than 90mm Hg. This two-year assessment was performed by independent field investigators, who visited households of all participants and measured their blood pressures. Multiple mop-up visits were conducted, and facilitation by ASHAs and study supervisors was solicited to improve proportion of follow-ups.

### Statistical Analysis

ASHAs and study supervisors entered all study-variables obtained in baseline survey, confirmatory-visits, UPHC-visits, or on follow-up using mobile-phone based data collection system. All the variables were stored on cloud-based CommCare data-management system. The demographic and cardiovascular-risk variables (age, gender, education level, wealth quintiles, tobacco use, alcohol consumption, physical activity levels, body-mass index, waist circumference) were obtained from entries in the baseline questionnaire. History of hypertension obtained at baseline and subsequently obtained SBP and DBP values were used for classification of hypertension status. Primary-care physician consultation decisions culminating in drug escalation, de-escalation or no action, were taken from UPHC visit logs. The distribution of these explanatory variables was compared with respect to outcome, using appropriate tests of significance. The difference was considered as significant if p-value for null hypothesis was less than 0.05.

We performed a multivariable logistic regression, to evaluate independent risk factors of uncontrolled hypertension. All explanatory variables that were marginally significant on univariate analysis (p<0.25) were included in the full model. Assumptions for using logistic regression were tested and Hosmer-Lemeshow Goodness-of-fit was also used. All data analysis and visualizations were done using Statistical software R (14) and *arsenal*, (15) *finalfit*, (16) *ggplot2*, (17) *ggpubr*, (18) *ggstatplot*,(19) *gtsummary*, (20) *performance* (21) and *tidyverse* (22) packages.

## Results

In November 2017 we initiated a baseline survey in 16 urban slum clusters of Bhopal. Of 6174 individuals surveyed at baseline, we identified a total of 1571 individuals (25.4%) with hypertension; and all of these were sought to be visited by CHWs once in every two months. A total of 1177 (74.9%) individuals completed two-year follow up visit done in November-December 2019. Individuals who were lost to follow up were more likely to be men, who were older, and were newly detected with hypertension. (Table S1) Of 1177 individuals who completed follow up, 623 (53%) knew about their hypertension status at baseline, and 554 (47%) were newly detected during screening. At baseline, 301 (26%) individuals were optimally controlled, and at the end of two years their proportion increased to 442 (38%), an absolute increase of 12% (95%CI 10.2-13.9). When we split these by status of hypertension diagnosis, of the 623 previously known hypertensives, 48% were optimally controlled at baseline, which became 46% at the end of two years, and of the 554 newly detected hypertensives, and 29% were optimally controlled at the end of two years. There was a significant decline of 1.7 (95%CI -2.9 to -0.5) and 1.7mm Hg (95%CI (−2.4 to -1.1) in mean SBP and DBP respectively from baseline to follow up. This change can be largely attributed to reduction in SBP among those with uncontrolled hypertension. (Table 1, Figure 1)

**Table-1:** Change in cardiovascular risk factors over two-year follow-up in the cohort (n=1177)

| Variables | Baseline (2017) | Follow Up (2019) | Absolute Difference (CI) | p-value |
| --- | --- | --- | --- | --- |
| **All participants (N=1177)** |  |  |  |  |
| SBP (mean, SD) | 146.6 (20.6) | 145.0 (21.2) | -1.7 (-2.9 to -0.5) | 0.007 |
| DBP(mean, SD) | 88.2 (12.6) | 86.5 (12.6) | -1.7 (-2.4 to -1.1) | <0.001 |
| Optimal BP control | 301 (26%) | 442 (38%) | +141 (12%) | <0.001 |
| BMI(mean, SD) | 25.7 (4.9) | 25.8 (5.2) | +0.1 (-0.05 to 0.18) | 0.257 |
| Obese or overweight | 536 (54%) | 533 (54%) | -3 (1%) | 1.000 |
| Waist Circum. (mean, SD) | 89.4 (14.4) | 92.2 (12.2) | 2.9 (2.3 to 3.5) | <0.001 |
| Men | 89.4 (19.5) | 92.5 (11.5) | 3.2 (1.1 to 5.2) | 0.002 |
| Women | 88.6 (16.8) | 92.1 (12.7) | 3.4 (1.9 to 5.0) | <0.001 |
| Abdominal Obesity | 817 (79%) | 868 (82%) | 51 (3%) | 0.0008 |
| **Previously known hypertension at baseline (N=623)** |  |  |  |  |
| SBP (mean, SD) | 141.8 (22.9) | 141.6 (21.0) | -0.2 (-1.9 to 1.5) | 0.825 |
| DBP(mean, SD) | 84.3 (13.4) | 83.8 (12.3) | -0.5 (-1.4 to 0.5) | 0.316 |
| Optimal BP Control | 301 (48%) | 284 (46%) | -17 (2%) | 0.277 |
| BMI(mean, SD) | 26.2 (4.9) | 26.3 (5.0) | 0.1 (-0.05 to 0.26) | 0.178 |
| Obese or overweight | 320 (59%) | 313 (57%) | -7 (2%) | 0.371 |
| Waist Circum. (mean, SD) | 89.9 (14.8) | 93.1 (12.3) | 3.2 (2.3 to 4.0) | <0.001 |
| Men | 89.7 (16.3) | 93.8 (11.8) | 4.0 (1.2 to 6.9) | 0.005 |
| Women | 89.2 (18.5) | 92.7 (12.4) | 3.6 (1.4 to 5.7) | 0.001 |
| Abdominal Obesity | 464 (85%) | 488 (87%) | +22 (2%) | 0.021 |
| **Newly detected hypertension at baseline (N=554)** |  |  |  |  |
| SBP (mean, SD) | 152.0 (16.1) | 148.7 (20.7) | -3.3 (-5.0 to -1.6) | <0.001 |
| DBP(mean, SD) | 92.7 (10.1) | 89.5 (12.3) | -3.2 (-4.14 to -2.2) | <0.001 |
| Optimal BP control | 0 (0%) | 158 (29%) | +158 (29%) | <0.001 |
| BMI(mean, SD) | 25.2 (4.9) | 25.2 (5.4) | 0.0 (-0.15 to 0.18) | 0.865 |
| Obese or overweight | 216 (48%) | 220 (49%) | +4(1%) | 0.429 |
| Waist Circum. (mean, SD) | 88.7 (14.0) | 91.3 (12.0) | 2.6 (1.7 to 3.5) | <0.001 |
| Men | 89.1 (21.4) | 91.7 (11.2) | 2.6 (0.26 to 5.4) | 0.075 |
| Women | 87.8 (13.8) | 91.0 (13.0) | 3.2 (1.0 to 5.5) | 0.005 |
| Abdominal Obesity | 353 (72%) | 380 (76%) | +27(4%) | 0.161 |

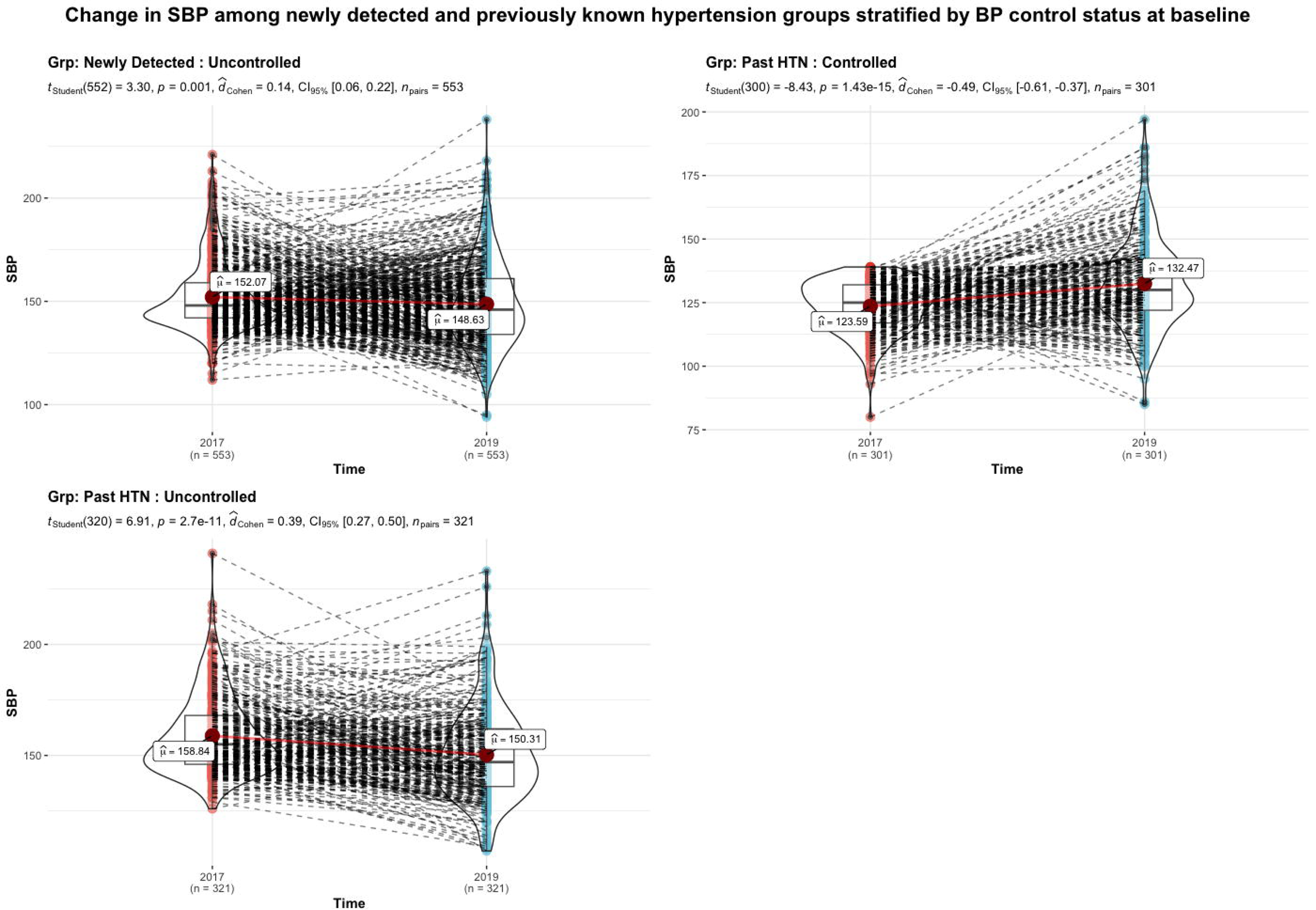

The proportion of individuals with uncontrolled blood pressure was progressively higher in each age band (55%, 63%, 67% and 73% in those with baseline age of 30-44, 45-59, 60-74, and above 75 years respectively). We observed a significant decline in SBP and DBP, across all age-bands of individuals with uncontrolled hypertension at baseline. About 65% of individuals who were sedentary at baseline, developed uncontrolled hypertension, as compared to only 54% of those who were non-sedentary i.e. physically active. This difference was statistically significant. Individuals who were newly detected to have hypertension, and those who had elevated blood-pressure at baseline, were more likely to be uncontrolled on follow-up. (Table 2, Figure 2)

**Table-2:**
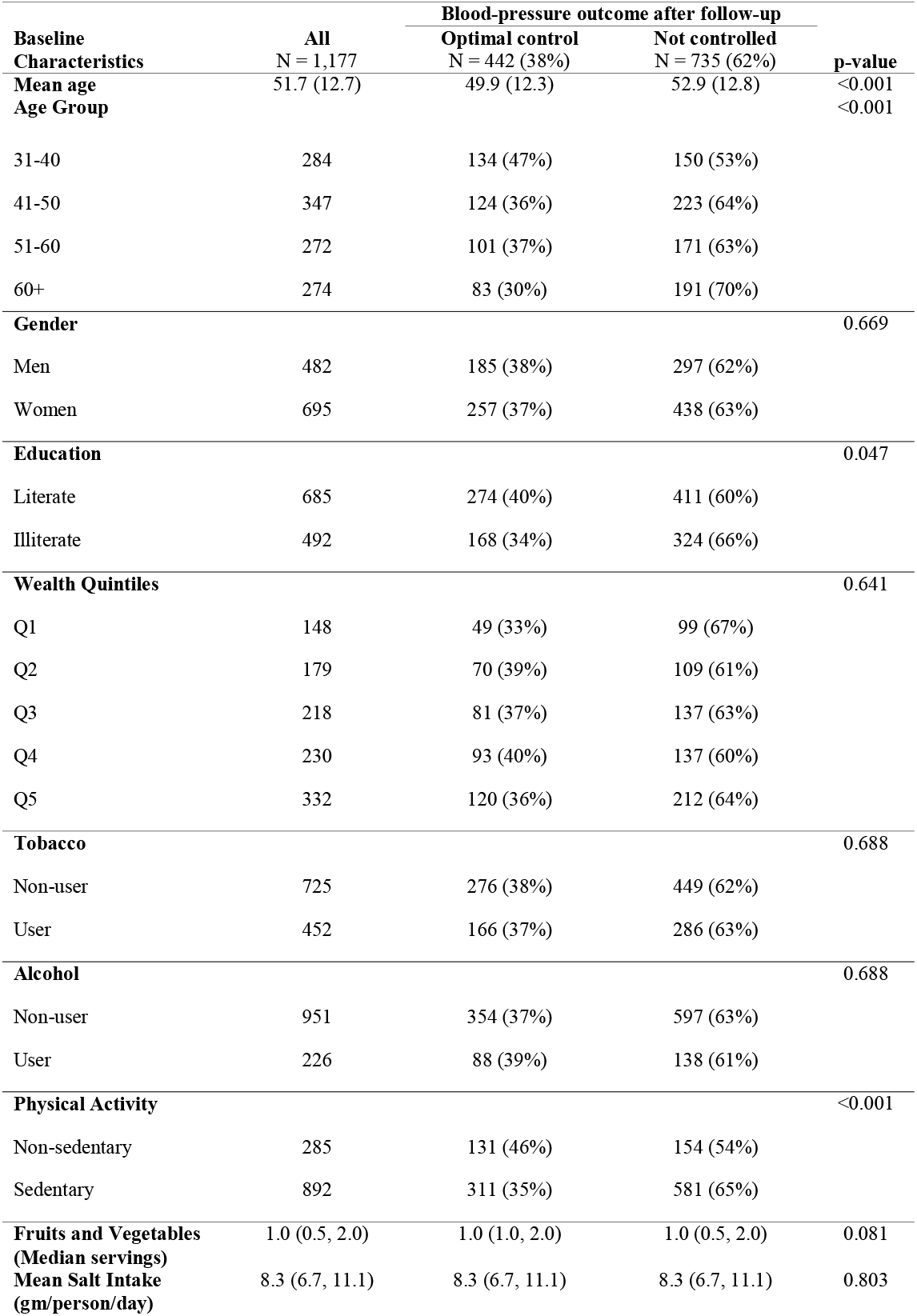
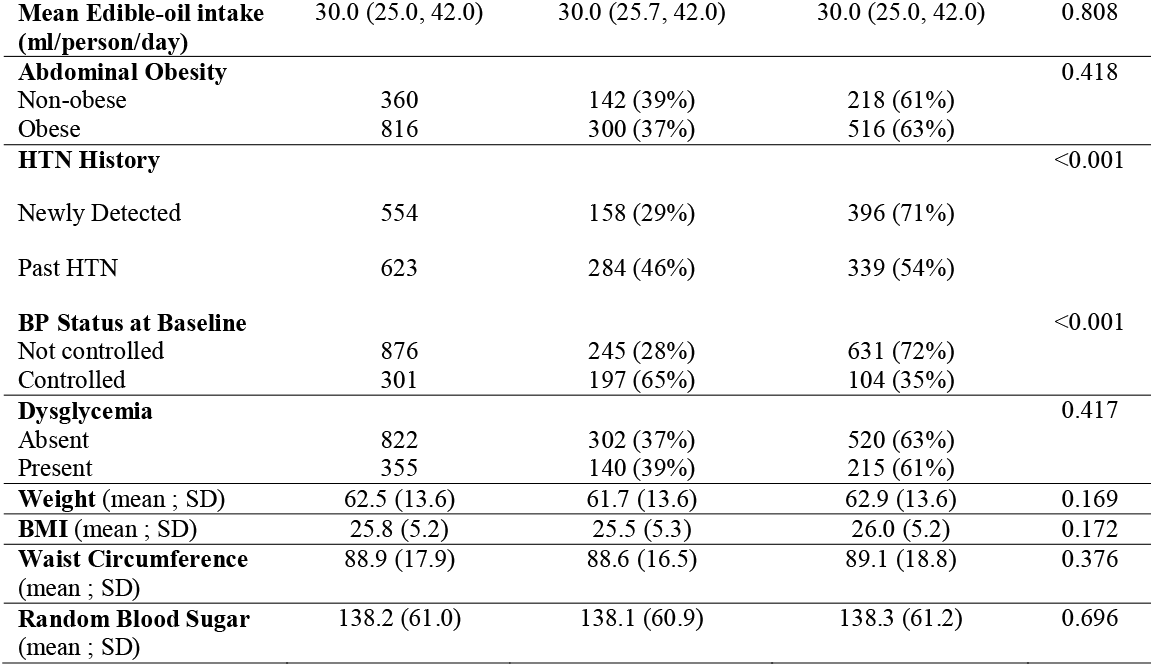
Distribution of status of BP control among various socio-demographic, behavioral and biological variables.

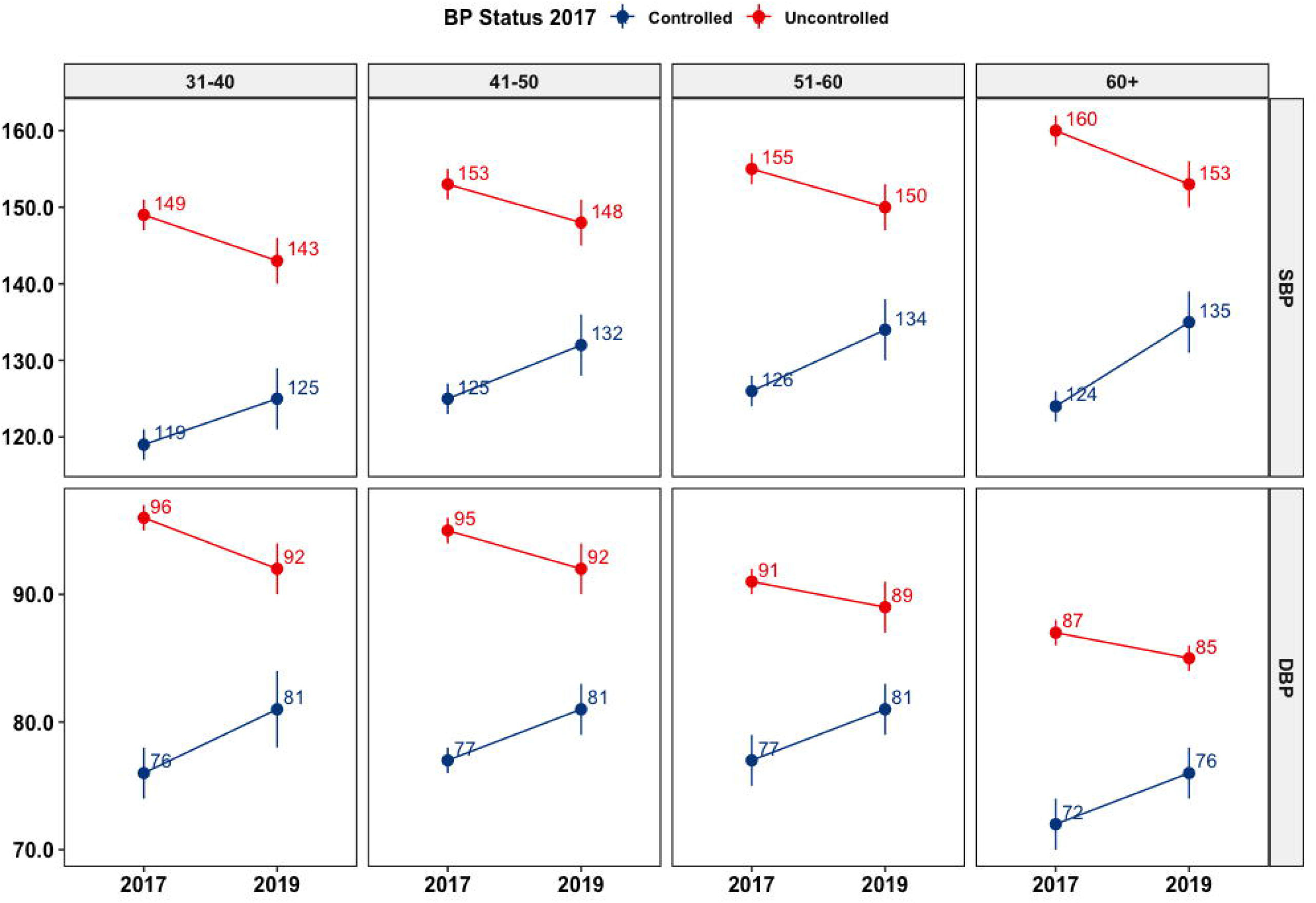

A total of 699 (59.4%) individuals reported to be on anti-hypertensive drug therapy, 533 of these were linked to UPHC, and 166 to private-care providers. All these individuals received follow up visits by ASHAs, and during these visits self-reported adherence to prescribed therapies was similar in both optimally controlled and uncontrolled individuals. (84.4% vs 85.8%; p=0.842) An independent adherence assessment was also done at the end of one-year in 263 of the UPHC attendees, and perceived adherence was lower in those subsequently found to be uncontrolled (69.5% vs 79.1%; p=0.006). The proportion of individuals who were uncontrolled after two years was significantly higher amongst those who were linked to UPHC as compared to those linked to private-providers (67% vs 61%; p=0.009). Of those linked to UPHC, median number of visits to the facility was similar in optimally controlled and uncontrolled subgroups (p=0.436) and 116 (21.7%) individuals had one or more visit culminating in a drug-escalation advise. While 24 (20.6%) of individuals who were advised drug-escalation were optimally controlled, remaining 92(79%) were not. Of the 478 of 1177 individuals (40.6%), defaulted for initiation of drug-therapy, and 276 (58%) had their blood pressures above 140/90 mm Hg at two-year follow up.

We performed multivariable logistic regression to identify risk-factors for uncontrolled hypertension. Hosmer-Lemeshow *goodness-of-fit* test was non-significant, there was no multi-collinearity, assumptions of homogeneity of variance, residual distribution and absence of influential observations were met, however model’s explanatory power was weak (Tjur’s R2=0.06, RMSE=0.46). After adjusting for age, and gender newly detected hypertension (OR 2.42 (1.78-3.31)), and BMI (OR 1.04 (1.01-1.07)) were significant independent risk-factors for uncontrolled hypertension. (Table 3)

**Table-3:**
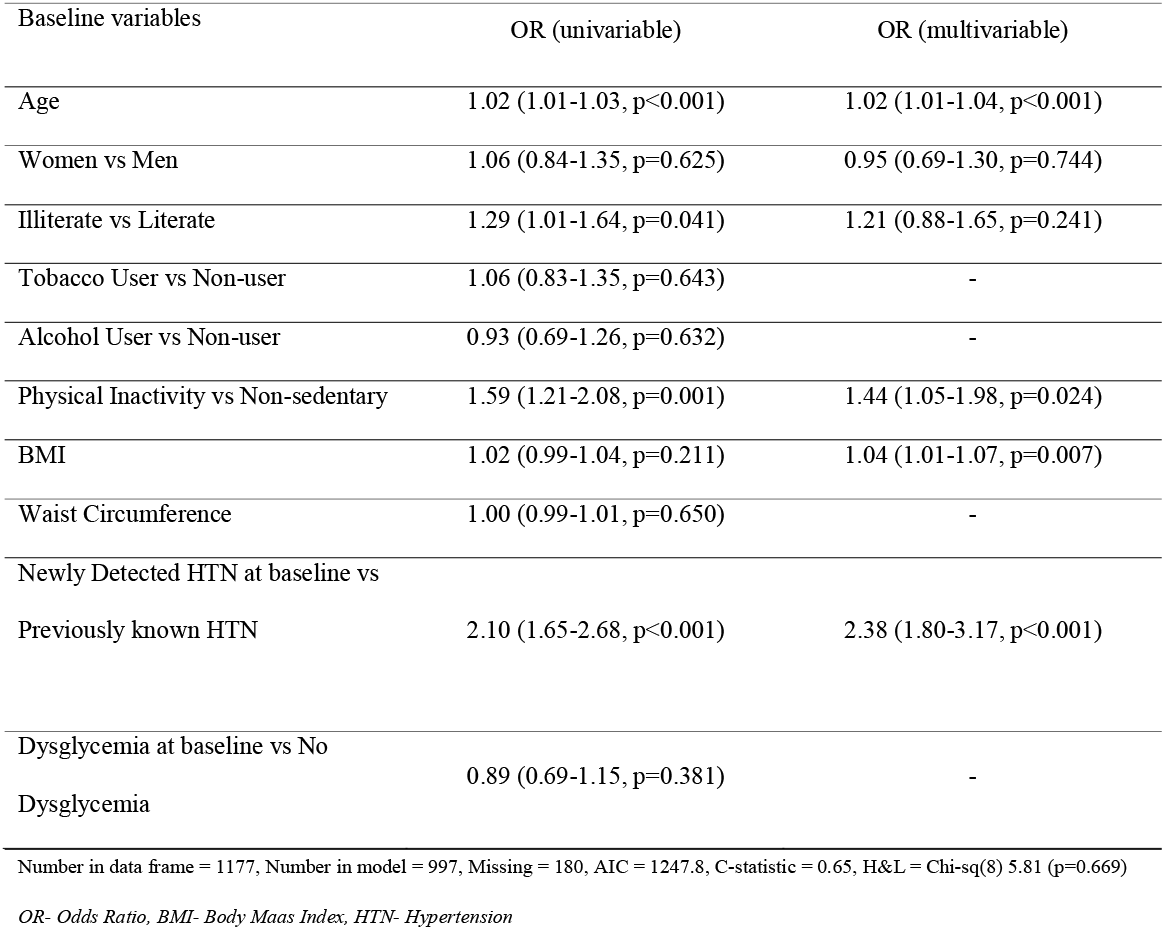
Logistic regression analysis for determinants of uncontrolled hypertension.

## Discussion

In the current study we found that after initial screening and two-year follow-up, about six of every ten individuals with hypertension were on-treatment, and about four were optimally controlled. While overall proportion of individuals with optimally controlled blood-pressure rose over the two-year period, this modest increase was due to blood-pressure control in less than a-third of newly detected hypertensives. More than two-thirds of the remaining newly detected and about half of previously known hypertensives remained uncontrolled. Those who were uncontrolled, were more likely to be older, had a higher BMI, and were newly detected to have hypertension. These findings demonstrate that obtaining blood-pressure control in vulnerable communities, though feasible is challenging. These findings provide a benchmark for NPCDCS, in terms of achievable goals within short periods of follow-up.

A recent systematic review (23) reported that in India only 13% of all individuals with hypertension are treated, and a meagre eight percent are controlled. Our baseline screening bridged the awareness gap, and CHW based adherence reinforcement improved proportion of on-treatment and controlled individuals from 43% and 29% to 60% and 38% respectively. While these numbers are encouraging, and approach and even exceed proportions for control achieved in previous studies in high-income settings,(24,25) they bring-forth challenges of initiating anti-hypertensive drugs in all who need them, improving adherence in those initiated, and appropriate dosing requirements.(26) In the current study education, wealth, or gender did not increase risk of uncontrolled hypertension. These socioeconomic characteristics are perceived as proxy-indicators for awareness, access or empowerment. It is likely that more complex behavioural phenomenon such as fear, reluctance and adverse impact of being labelled with an illness, affected both initiation and adherence to prescribed drug therapies.(27) We found that risk of being uncontrolled was consistently higher with increasing age, a factor that contributes to therapeutic inertia.(28) Further, presence of fixed-dose combinations in the drug formulary in UPHCs could have helped reduce inertia,(29) as use of combination treatment in newly detected hypertensives is likely to achieve a faster blood-pressure control.(24)

Similar findings were reported in recent studies on predictors of hypertension control in primary health care settings.(30–32) These studies have reported age, gender, race, Waist-Hip Ratio, BMI, presence of diabetes mellitus, intake of multiple drugs and lack of knowledge as predictors. (30–32) Physical activity is important to achieve blood-pressure control, and is an important component of various population based programs.(33) Self-reported physical activity levels correlate well with reduced blood pressure levels.(34) In the current study, hypertensive individuals who reported themselves to be sedentary at baseline were more-likely to be uncontrolled two years later. While this effect observed on univariate analysis, was not significant when adjusted for age and gender, it is an important life-style measure that has a strong legacy effect. Improved physical activity achieves blood pressure reduction through diverse mechanisms such as reduction in vascular resistance, sympathetic activity, inflammation and stress.(34) It also leads to lower BMI, which was an independent predictor of blood-pressure control in our study. While some studies, such as SPRINT trial have found no relationship between BMI and blood-pressure control, (35) another large study from China found a significant lowering effect.(36) While mean BMI of participants in SPRINT trial was 29 kg/m2, it was 24.4 kg/m2 in the Chinese study. Mean BMI of our participants was 25.8 kg/m2, closer to that of the Chinese study. It is likely that blood-pressure control in population sub-groups with a lower BMI is more feasible, as compared to those with obesity.

Improving blood pressure control is challenging, and regular follow up with intensive pharmacotherapy is most useful to achieve this goal. In a Cochrane review of 56 randomized control trials, other measures such as self-monitoring, appointment reminders, and health-education had little independent effects and were less useful.(37) Feasibility of achieving even lower targets than envisaged in NPCDCS, was demonstrated in intensive blood-pressure control arm in SPRINT trial.(38) Similar intensive management is also possible through CHWs in low-income settings. Two recent trials have demonstrated this effect. First, HOPE4 study demonstrated that CHW based intensive blood pressure control can improve proportion of optimally controlled individuals to 69% as compared to 30% in standard care arm.(39) Second, in another randomized control trial from Argentina, CHW led multi-dimensional care that involved intensive health-coaching, home BP monitoring, audits and feedbacks optimally controlled 72% individuals.(9) When CHWs are provided with intensive supportive supervision, empowered to obtain quick feedbacks on treatment decisions, it can help improve therapeutic inertia and achieve better control levels.(40)

## Conclusion

We have established a cohort of individuals with hypertension, with an aim to improve CHW based care delivery in vulnerable populations and generate evidence that may help implementation of NPCDCS. We used efficient data-management systems, within the programmatic settings to establish a system of community-based follow-up. Our study however has had some limitations. We could follow up only three-fourths of our intended cohort, mostly due to migration, and limited availability at home due to long working hours of many participants. Proportion of individuals on-treatment was modest, reflecting the real-world apprehensions many individuals with hypertension have, regarding initiation of drug therapy. Further, the process of obtaining prescription refills from a health-facility often requires missing work on that day, which may not be feasible for the working populations. Last, our follow up duration was only two years. While this is short, but we believe it is sufficient to demonstrate improvements within program settings.

To conclude, in the current study we found about six of every ten individuals with hypertension were on-treatment, and about four of them were optimally controlled. Our study has implications for implementation and practice of NPCDCS. These findings truly provide a benchmark for NPCDCS which are in terms of achievable goals within short periods of follow-up. Newly detected individuals with hypertension, and those who are obese, are two such subgroups that need more intensive monitoring and follow-up to improve optimal control levels in the community. CHWs based follow-ups are feasible, but more intensive treatments and follow-up systems are needed for better blood-pressure control outcomes.

**What was already known?**

In India only around 13% of all individuals with hypertension were on anti-hypertensive medications, and a meagre eight percent were controlled. NPCDCS program is rolled out to improve this situation.

**What this study adds?**

In the current study we found that after initial screening and two-year follow-up, about six of every ten individuals with hypertension were on-treatment, and about four were optimally controlled.

Those who were uncontrolled, were more likely to be older, had a higher BMI, and were newly detected to have hypertension.

These findings demonstrate that obtaining blood-pressure control in vulnerable communities, though feasible is challenging.

## Data Availability

Raw data of this study is not deposited in any public repository. However, anonymized raw data of this study would be available to academicians or researchers on reasonable request to corresponding author.

## Acknowledgement

Investigators would like acknowledge efforts of all Community Health Workers (ASHAs) and their officials affiliated to Urban Public Health system in implementing this study.

